# Prevalence of Diabetes Mellitus Type-2 among HCV Patients Admitted to The Virology Unit at Fever Hospital in Ismailia and Assessment of The Credibility of HBA1C in Assessing Diabetic Control among Diabetic Patients

**DOI:** 10.1101/2022.08.31.22279437

**Authors:** Mohamed Khaled Mohamed Ibrahim, Sara Elsayed, Nashwa Mostafa, Aya Ahmed, Khadiga Ahmed Abdelrehem, Afnan Mahmoud, Yasser Leil, Alzahraa Mohamed Adel, Abdelrahman Elsayed, Ahmed Abdelgawad, Mayar Mohamed, AbdAllah El-bally

**Author notes:** Contact Information (Chairman of the group): Name: Mohamed Khaled Mohamed Ibrahim. Prof. Abdelraouf Eldeeb Professor of internal medicine.

## Abstract

**Background:** Glucose is the main source of energy for the human body. The liver plays an important role in physiological glycemic control as it produces, stores & release glucose depending on our need for glucose through involvement in several glucose metabolism processes including glycogenesis & glycolysis.After meal, carbohydrates in the food we eat are reduced into simplest form, glucose. Excess glucose removed from body and converted into glycogen in a process called glycogenesis.

Many studies approved that type 2 diabetes and hepatogenous diabetes are associated with increased risk of complication of chronic liver diseases and mortalityGenetic factors play a major role as well. HCV is considered a diabetogenic agent through multiple mechanisms: autoimmune phenomena, direct cytotoxic effect on pancreatic cells, and, blockage of insulin receptors at cellular levels. Alcoholic chronic liver disease affect both hepatocytes and pancreatic islet cells. Diagnosis of hepatogenous diabetes may be difficult as clinical manifestation in early stages of chronic liver disease may be absent and fasting plasma glucose may be normal.So in our study we want to identify best investigation to assess diabetes in chronic liver disease patients. Three prospective studies were collected assessing impact of diabetes among chronic liver disease patients; mainly the outcome, and all of them demonstrate lower 5-year cumulative survival

**Aim:** To provide data to augment the standard of care in diabetic patients with chronic liver disease..

**Methods:** Through a formal permission and access to the (VF-IFH) data and recording system.Data are recorded in a paper-based database system. Collection of data will be via copying the data into an excel sheet. Diabetic status; fasting glucose will be used as a gold standard to divide patients into diabetics (abnormal fasting glucose) and non-diabetics (normal fasting glucose level). Assessment of HbA1C values and patients’ diabetic control, as documented in (VF-IFH) database the sample size for this study is 167 of chronic hepatitis C patients

**Results:** The sample size for this study is 167 of chronic hepatitis C patients. Out of 167 questioned patients, 30.54% are diabetics. 25.5% of diabetic patients have normal HA1C (controlled) & 74.5% have abnormal HA1C (uncontrolled). 78.43% of patient have elevated fasting plasma glucose & about 21.57% have normal values.About 56.86% of hepatitis C patients that have diabetes, have abnormal kidney function (elevated serum creatinine).

**Conclusion:** Chronic liver disease affects glucose metabolism, ranging from mere glucose intolerance to overt diabetes, which is known as hepatogenous diabetes.We find that, about 50, 54% of chronic hepatitis C patients are diabetics with 25, 5% have normal HA1C, 74, 5% have abnormal levels. With no limitations, results precisely answer our question, demonstrate that hepatogenous diabetes is a common problem among chronic liver patients and HA1C is not a standard assessment tool for diabetes.Finally, we wait more researches to explain the pathological basis of the mysterious relation between cirrhosis and HA1C.

## INTRODUCTION and LITTERATURE REVIEW

Glucose is the main source of energy for the human body. The liver plays an important role in physiological glycemic control as it produces, stores & release glucose depending on our need for glucose through involvement in several glucose metabolism processes including glycogenesis & glycolysis.

After meal, carbohydrates in the food we eat are reduced into simplest form, glucose. Excess glucose removed from body and converted into glycogen in a process called glycogenesis. ^1^

During fasting or when blood glucose level declines, hepatic cells convert glycogen into glucose and release them into blood till level of glucose approaches normal in a process called glycogenolysis.

In chronic liver disease: there is progressive destruction of liver parenchyma over a period greater than 6 months leading to fibrosis and cirrhosis. Due to progressive loss of hepatocyte function, glucose metabolism is impaired in a process called hepatogenous diabetes.

What is hepatogenous diabetes? DM which develops as a complication of cirrhosis is known as Hepatogenous Diabetes^2^

Patient with CLD may have two types of diabetes; type 2 DM: related to metabolic syndromes that cause non-alcoholic fatty liver disease, hepatocellular carcinoma. Hepatogenous Diabetes that result as a result of liver cirrhosis.

Many studies approved that type 2 diabetes and hepatogenous diabetes are associated with increased risk of complication of chronic lver diseases and mortality. ^3^

Pathophysiology of hepatogenous diabetes is complex and not clearly understood ^2^. Hepatogenous diabetes may result from; Decrease extraction of insulin by damaged liver. And liver dysfunction itself has a toxic effect on pancreatic islets and this proved when hepatogenous diabetes improves after successful liver transplantation. ^3^ As well as insulin Resistance of peripheral tissues (muscles, liver, fat) one of the major mechanism involved in pathophysiology. Genetic factors play a major role as well. HCV is considered a diabetogenic agent through multiple mechanisms: autoimmune phenomena, direct cytotoxic effect on pancreatic cells, and, blockage of insulin receptors at cellular levels. Alcoholic chronic liver disease affect both hepatocytes and pancreatic islet cells.

Diagnosis of hepatogenous diabetes may be difficult as clinical manifestation in early stages of chronic liver disease may be absent and fasting plasma glucose may be normal.

So in our study we wanted to identify best investigation to assess diabetes in chronic liver disease patients. Three prospective studies were collected assessing impact of diabetes among chronic liver disease patients; mainly the outcome, and all of them demonstrate lower 5-year cumulative survival ^4,5^

## AIM OF THE STUDY

To provide data to augment the standard of care in diabetic patients with chronic liver disease.

## OBJECTIVE(S)

### Primary Objective(s)

1. To determine prevalence of type2 diabetes mellitus among patients with chronic liver disease.
2. To determine effect of chronic liver disease on credibility of glycosylated hemoglobin (Hb A1C) as a standard assessment tool for diabetes.

### Secondary Objective(s)

1. To assess prevalence of family history among diabetic patients with chronic liver disease.
2. To assess treatment modalities among diabetic patients with chronic liver disease.

## RESEARCH QUESTION

Does the prevalence of diabetes mellitus type 2 among HCV patients lies within the range 28 – 55 %?

Does chronic liver disease affect the credibility of HbA1C as a standard assessment tool for diabetes?

## RESEARCH HYPOTHESIS

The prevalence of diabetes mellitus type 2 among HCV patients lies within the range 28 – 55%.

Chronic liver disease affects the credibility of HbA1C as a standard assessment tool for diabetes

## SUBJECTS & METHODS

### STUDY DESIGN

Observational Analytic cross-sectional study.

### STUDY SETTING

Virology unit at Ismailia fever hospital (VF-IFH).

### STUDY POPULATION

Chronic HCV patients admitted to virology unit at Ismailia fever hospital (VF-IFH).

## SAMPLING

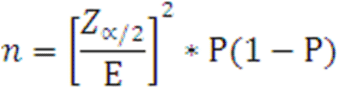

### Sample size

**n**= sample size

**Z α/2** = 1.96 (The critical value that divides the central 95% of the Z distribution from the 5% in the tail)

**p** = the prevalence of type 2 DM among HCV patients = 31.5 %

**E** = the margin of error (=width of confidence interval) = 0.05

**so n = 331 subject**

### Sampling technique

Sample size will be collected randomly from all recorded patients in the last six month.

## METHODS

### Data Collection Tools & procedures

Through a formal permission and access to the (VF-IFH) data and recording system.

Data are recorded in a paper-based database system. Collection of data will be via copying the data into an excel sheet.

- Diabetic status; fasting glucose will be used as a gold standard to divide patients into diabetics (abnormal fasting glucose) and non-diabetics (normal fasting glucose level).
- Assessment of HbA1C values and patients’ diabetic control, as documented in (VF-IFH) database and records and by using … questionnaire

## STUDY DURATION & TIMETABLE

- Stage 1, protocol writing ---- 1-2weeks
- Stage 1, review of literature ---- 1-2weeks
- Stage 2, data collection and data analysis ---- 1-2 weeks
- Stage 3, presentation and publication ----1-6 months

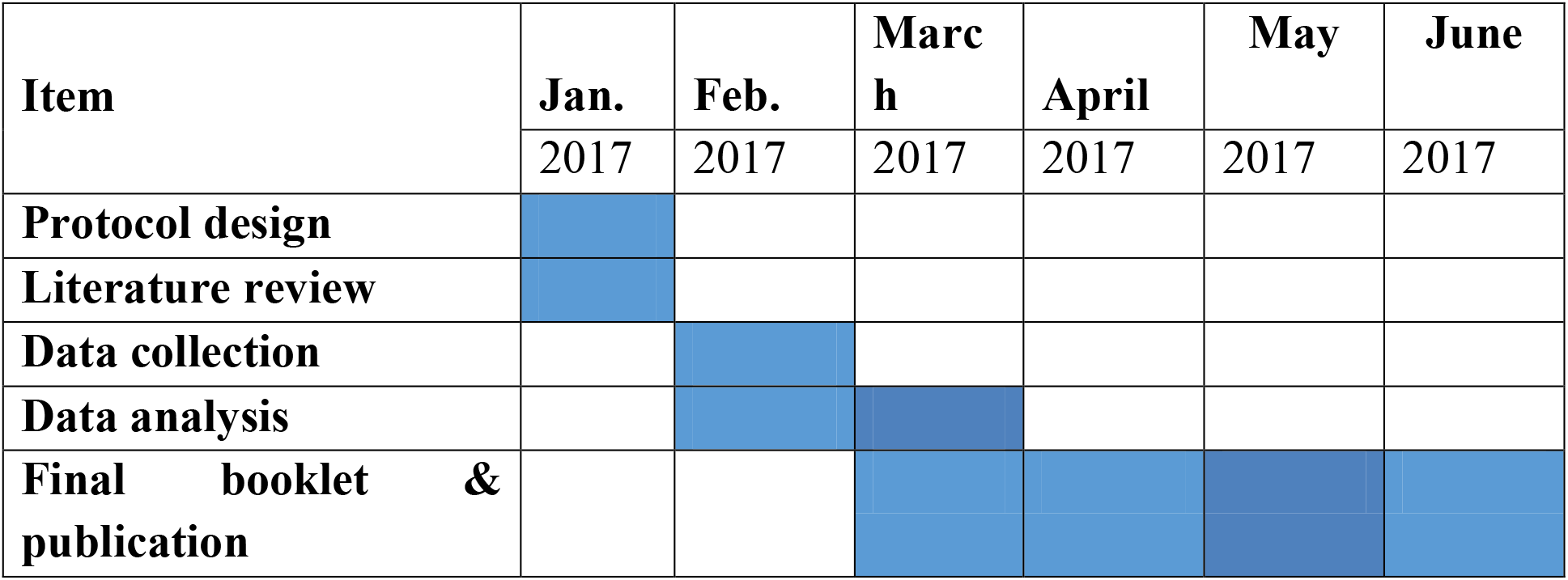

## STATISTICAL ANALYSIS PLAN

All statistical analyses were performed using the Statistical Package for Social Science (SPSS) version16. Data presentation was performed via table and graphs. Qualitative data was presents as numbers and percentages while quantitative data as mean ± Standard deviation. Kolmogorov test was be used to test normality. Parametric and non-parametric tests were used as required. Students (t) test was used for normally distributed. Chi square and Fisher’s exact tests would be used for qualitative variables. P value of <0.05 was considered statistically significant.

## ETHICAL CONSIDERATION

1. The project was be approval by the ethical committee of the Suez Canal University.
2. Permission of the managers of VF-IFH will be obtained.
3. Consent of the subjects included in the study without any obligation.
4. Participants were informed that responding is voluntary and that they can refuse responding without stating any reason.
5. Confidentially of the data collected as data was collected as codes without names.
6. Clear explanation of the study, questionnaire and the procedure to the subjects
7. No interference with work harmony.
8. A feedback about the results of the study was given to participants and their families.

## BUDGET

This study is self-funded by the authors.

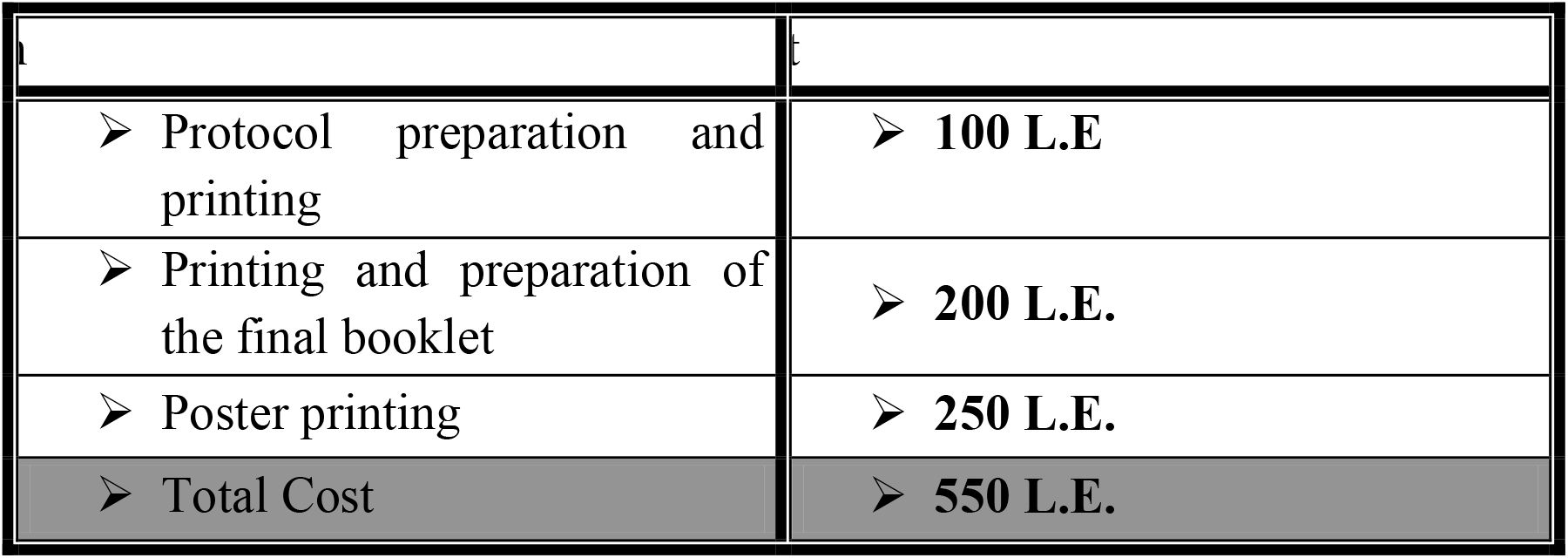

## Results

### Relation between HA1C values and serum creatinin levels

Using these data, we can also say that we cannot use HA1C to assess glycemic control or to predict complication in chronic hepatitis c patients with diabetes.

**Table 1:** Demograghic characteristics of chronic hepatitis C patients in studied population:

|  | variables | Frequency | Percent |
| --- | --- | --- | --- |
| **Age group (years)** | 20 – 40 =(0) | 26 | 15.66% |
|  | 40 – 60=(1) | 112 | 67.47% |
|  | 60 – 80=(2) | 27 | 16.27% |
|  | total | 166 | 100% |
| **Gender** | male | 67 | 40.72% |
|  | female | 99 | 59.28% |

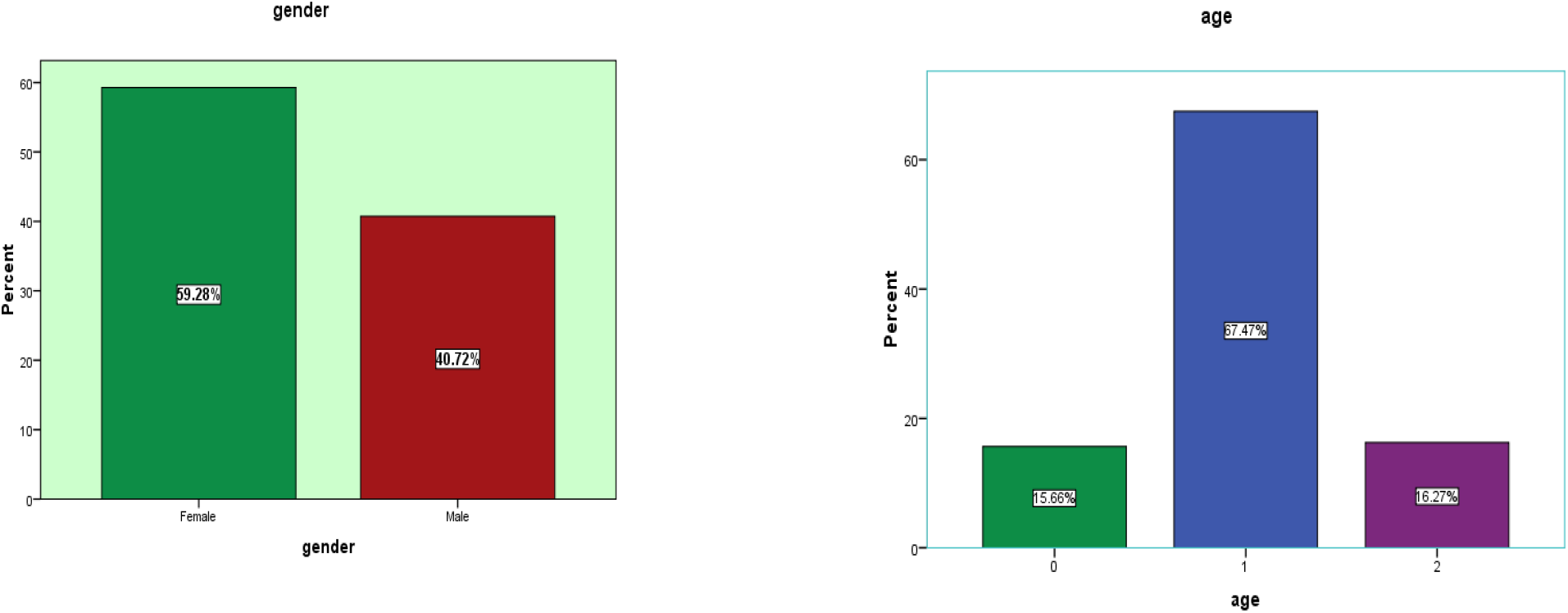

Our sample size was 166, 59.28% are female & 40.72% are males and most of them 112 patients lies between (40-60), 26 patients lies between (20-40) And 27 patients lies between (60-80).

**Table 2.**
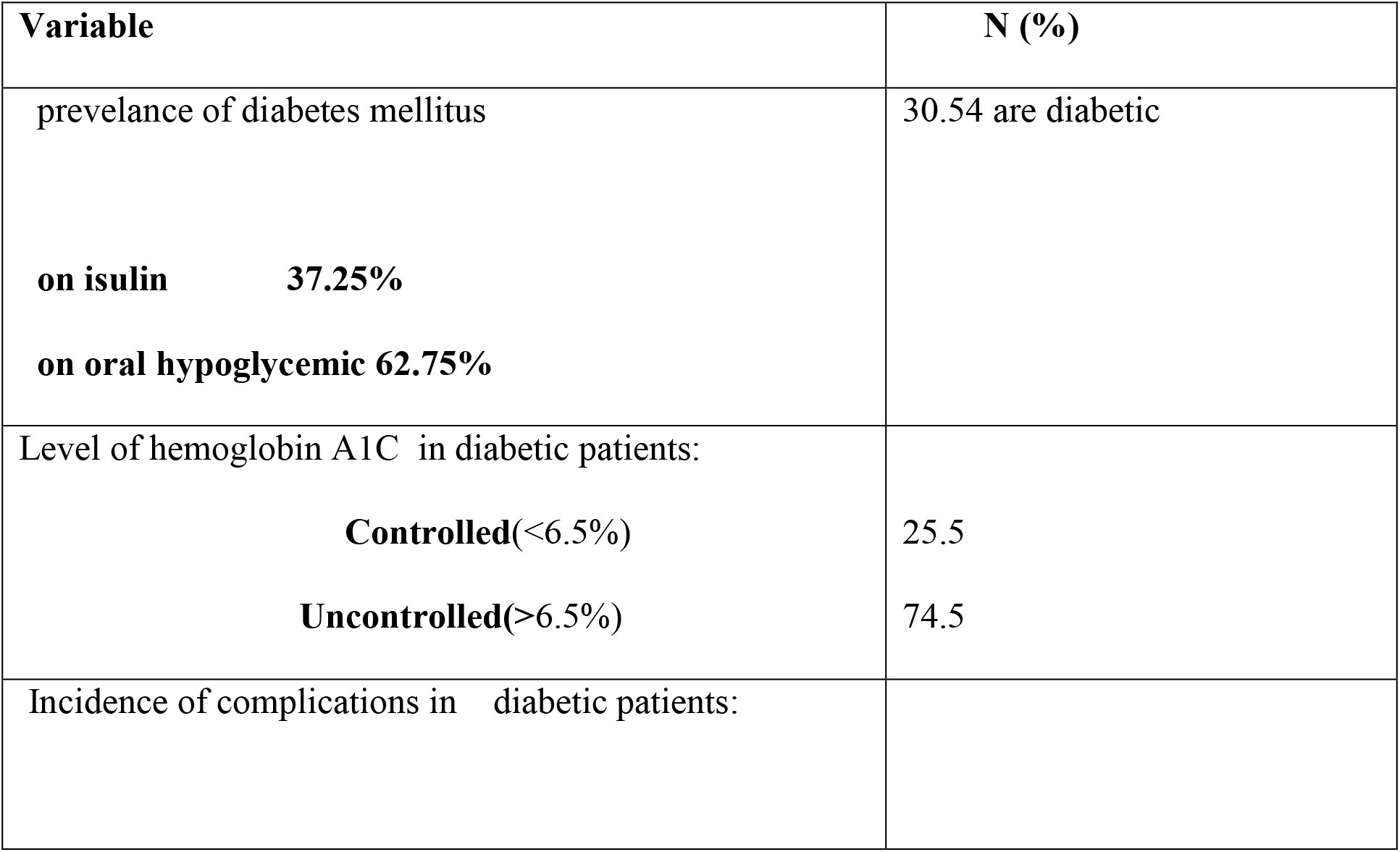
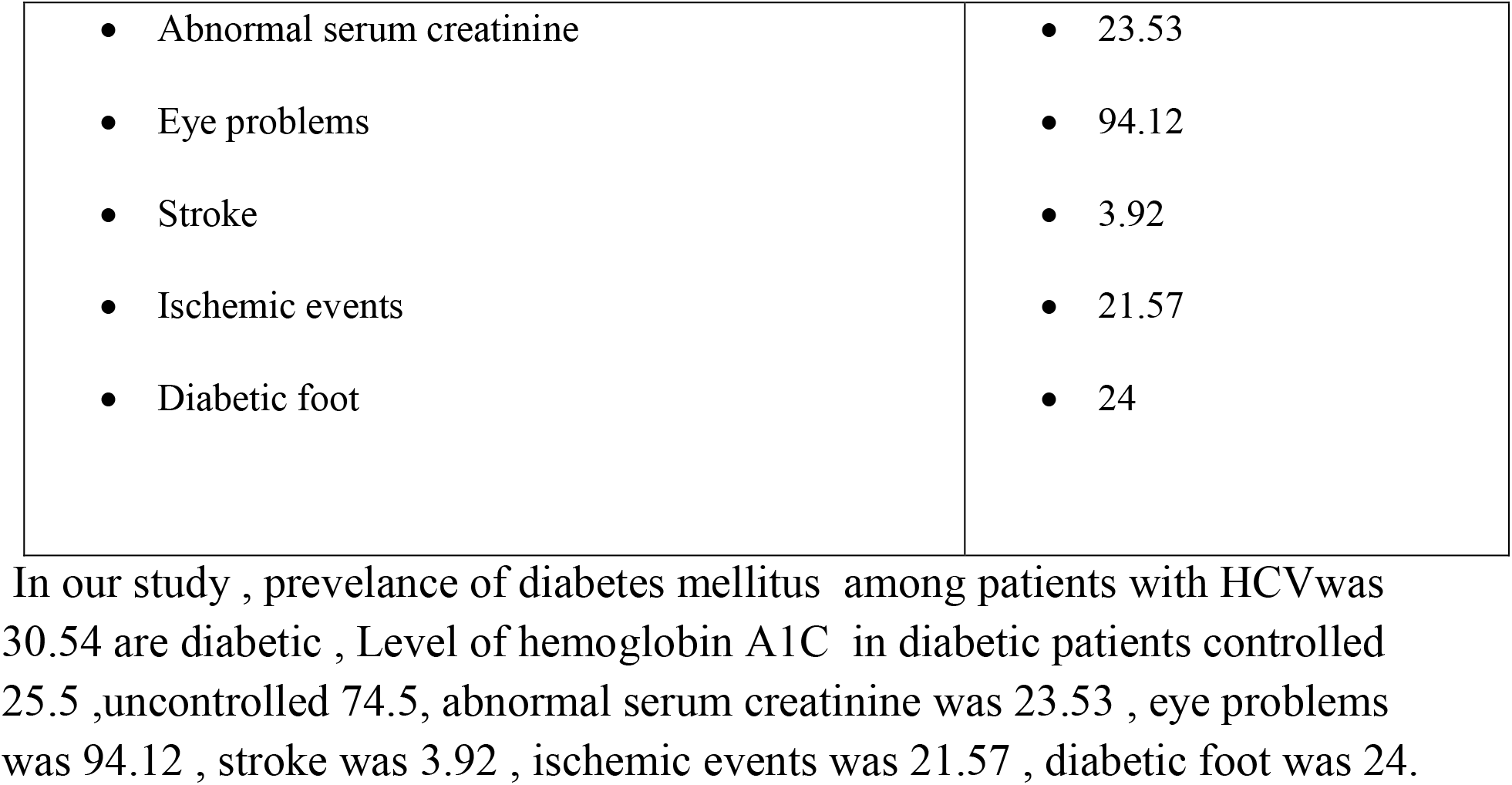
prevelance of DM, level of HBA1C and complications.

| Variable | N (%) |
| --- | --- |
| prevelance of diabetes mellitus | 30.54 are diabetic |
| on isulin 37.25% |  |
| on oral hypoglycemic 62.75% |  |
| Level of hemoglobin A1C in diabetic patients: |  |
| Controlled(<6.5%) | 25.5 |
| Uncontrolled(>6.5%) | 74.5 |
| Incidence of complications in diabetic patients: |  |
| • Abnormal serum creatinine<br>• Eye problems<br>• Stroke<br>• Ischemic events<br>• Diabetic foot | • 23.53<br>• 94.12<br>• 3.92<br>• 21.57<br>• 24 |

In our study , prevalence of diabetes mellitus among patients with HCV was 30.54 are diabetic , Level of hemoglobin A1C in diabetic patients controlled 25.5 , uncontrolled 74.5, abnormal serum creatinine was 23.53 , eye problems was 94.12 , stroke was 3.92 , ischemic events was 21.57 , diabetic foot was 24.

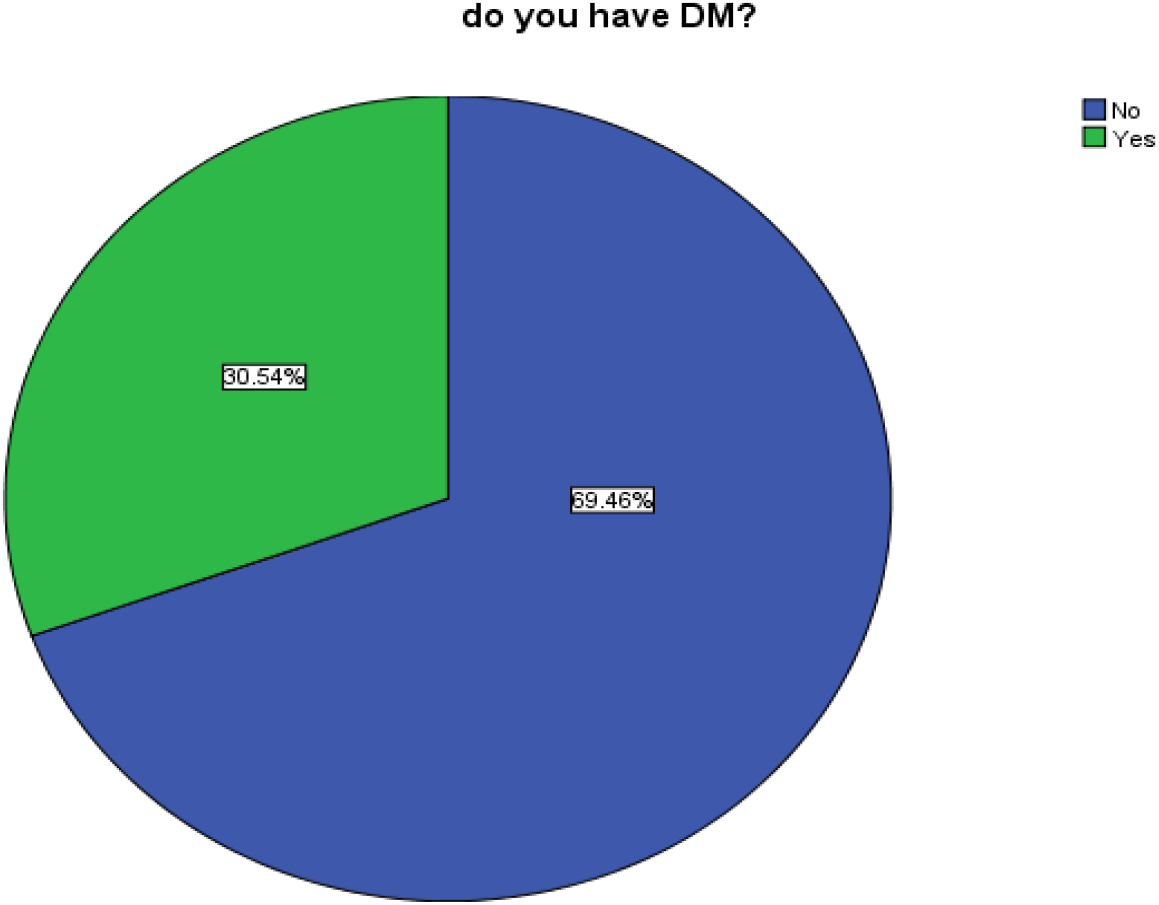

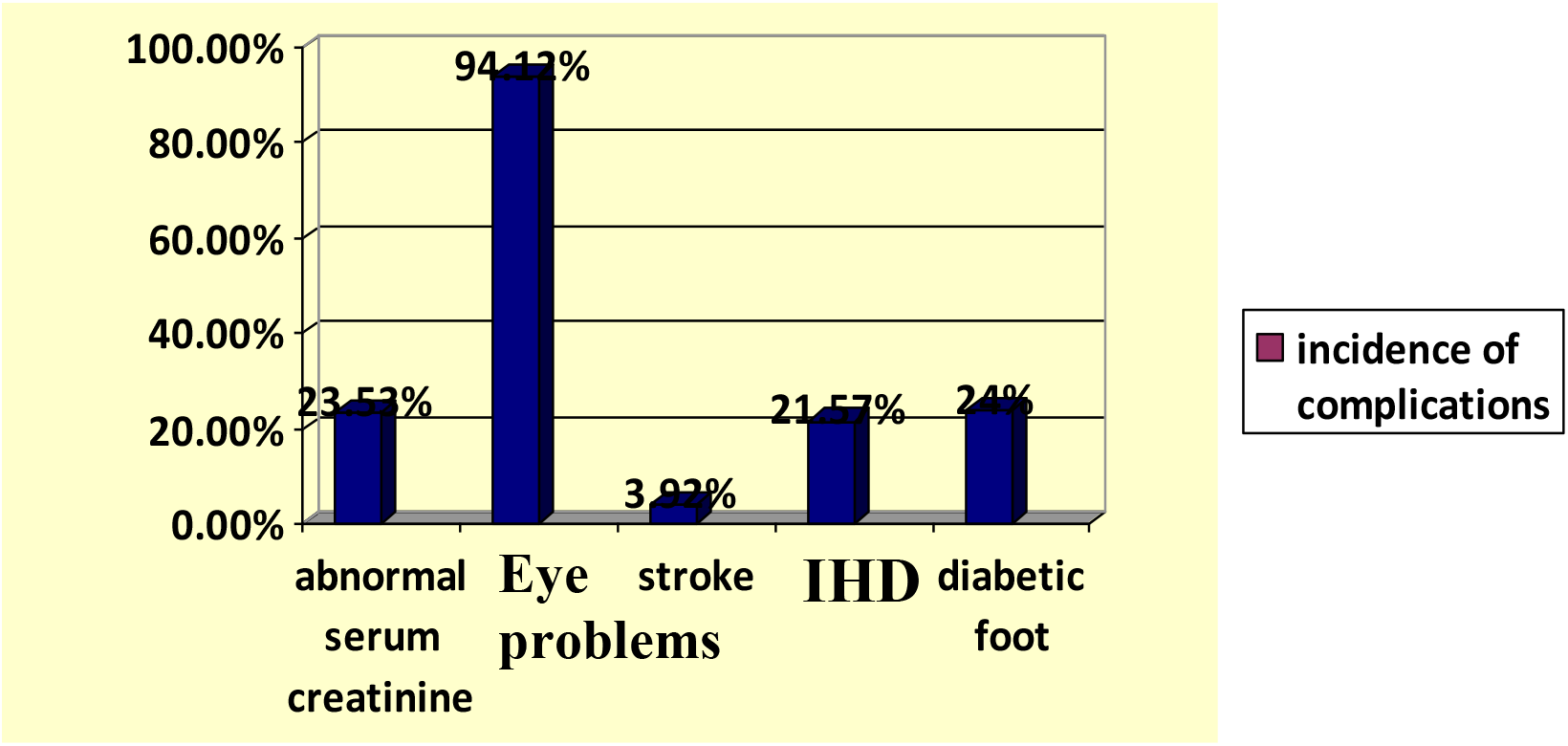

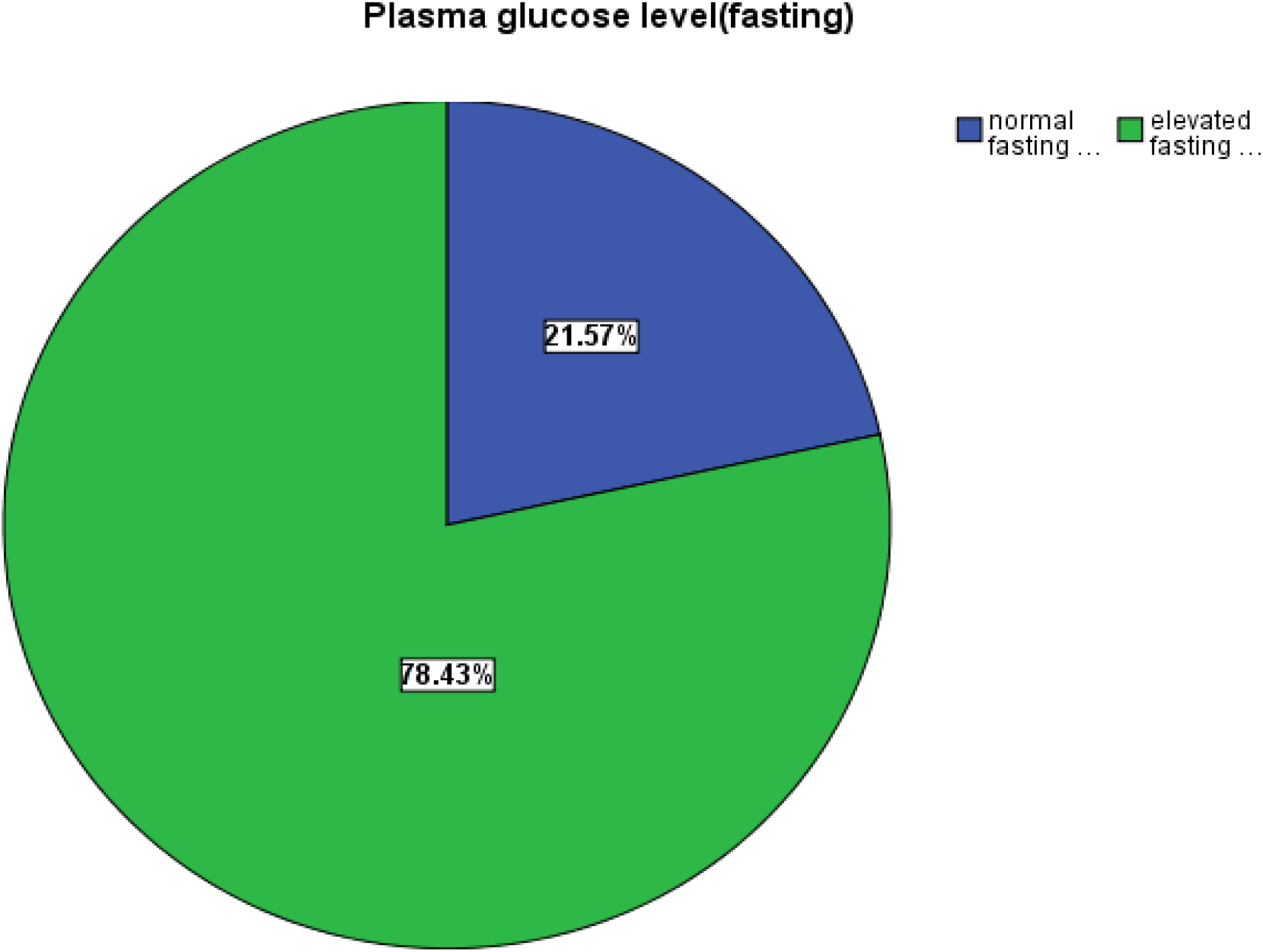

### Fasting plasma glucose

78.43% of patient have elevated fasting plasma glucose & about 21.57% have normal value

**Using chi-square test**, our **p-value** is 0.862*.there is no statistically significant assossiation between high plasma level & high levels of HA1C in chronic hepatitis C patients with diabetes.

**Table 4.** Association between plasma glucose level HA1C level (N = 51)

|  |  | HA1C level |  |  |  | Total |  | *p*-value |
| --- | --- | --- | --- | --- | --- | --- | --- | --- |
|  |  | uncontrolled |  | controlled |  |  |  |  |
| plasma glucose level | normal | 3 | 5.8% | 1 | 1.96% | 4 | 7.8% | .862* |
|  | elevated | 37 | 72.5% | 10 | 19.6% | 47 | 92.1% |  |
| Total |  | 40 | 78.3% | 11 | 21.5% | 51 | 100.0% |  |

\*. Statistically significant at p-value <0.05

## Discussion

Hemoglobin is the iron-containing oxygen transport metalloprotein in the red blood cells. Hemoglobin’s structure consists of a tetramer of two pairs of protein molecules: two α globin chains and two non–α globin chains.^1^ The α globin genes are HbA1 and HbA2, whereas the non–α globin genes include β, γ, δ. The normal adult hemoglobin molecule (HbA) consists of two α and two β chains (α_2_β_2_), and makes up about 97 % of most normal human adult hemoglobin.^2^ Other minor hemoglobin components may be formed by posttranslational modification of HbA. These include hemoglobins A1a, A1b, and A1c. Of these, A1c is the most abundant minor hemoglobin component. A1c is formed by the chemical condensation of hemoglobin and glucose which are both present in high concentrations in erythrocytes. This process occurs slowly and continuously over the life span of erythrocytes, which is 120 days on average. Furthermore, the rate of A1c formation is directly proportional to the average concentration of glucose within the erythrocyte during its lifespan.^2^ Hence, as levels of chronic hyperglycemia increase, so does the formation of A1c. This makes it an excellent marker of overall glycemic control during the time frame of the 120-day lifespan of a normal erythrocyte.

Results of the DCCT and UKPDS studies verified the close relationship between glycemic control measured by A1c and the risk for diabetes-related complications. A1c has been widely accepted as the standard used to measure glycemic control over the previous 3 month period and correlates with patients’ risk for developing diabetes-related complications.^3^ It is important to remember that A1c represents a weighted mean of glucose levels during the preceding 3 month time period. In other words, glucose levels during the most recent 6 week period will have a greater influence on the A1c result compared to levels from the prior 6 weeks.^4^ Thus, if the patient has experienced a recent acute change in glycemic control (i.e., treatment with glucocorticoids), the A1c value will be disproportionately affected by the most recent glucose levels.

So, Any condition that prolongs the life of the erythrocyte or is associated with decreased red cell turnover exposes the cell to glucose for a longer period of time, resulting in higher A1c levels. Similarly, any condition that shortens the life of the erythrocyte or is associated with increased red cell turnover shortens the exposure of the cell to glucose, resulting in lower A1c levels. Conditions such as acute and chronic blood loss, hemolytic anemia, and splenomegaly can all cause falsely lowered A1c results.^5^

### Liver cirrhosis (LC) is a glucose intolerance state

The liver performs numerous functions associated with glucose metabolism. Glucose uptake by the liver depends on circulating blood glucose concentration and contributes to the maintenance of glucose homeostasis. Glucose uptake by the liver is decreased in patients with liver cirrhosis because of a portal-systemic shunt and a decrease in viable hepatocytes, resulting in post-prandial hyperglycemia ^6^. Indeed, 80% of patients with liver cirrhosis also exhibit abnormal glucose tolerance and 25% have been diagnosed with diabetes ^7^ HbA1c measurements have limitations in particular diseases, including chronic liver diseases and liver cirrhosis. Hypersplenism in patients with liver cirrhosis results in a shorter half-life of erythrocytes, resulting in the underestimation of HbA1c ^8^. Therefore, HbA1c have been regarded as inadequate indicators of average blood glucose concentrations in patients with liver cirrhosis ^8^. However, these results derived from studies in which patients performed 7–8 self-monitoring blood glucose (SMBG) tests per day, with average glucose levels determined from individual, discontinuous glucose concentrations. Therefore, it remains unclear whether HbA1c is inappropriate indicator of average glucose levels in patients with liver cirrhosis.

In our study we tried to lightly put the HbA1C to the test by measuring it to the fasting blood glucose of the patient. Though it is not a perfect testing setting, but due to limited budget and resources that was the best modality to go around the test. But we still would like to raise fund to retest the HbA1C against more formidable and stronger markers especially the continuous glucose monitoring scales – to test credibility, and then work on a less invasive, more practical markers to use in the liver compromised patients as the mostly available-the HbA1C and fructosamine are compromised markers in these patients. And here’s a brief introduction to these markers.

### ALTERNATIVES TO USING A1C

In situations where A1c may not accurately reflect glycemic control, using an alternative index is desirable. Such potential indices include fructosamine, glycated albumin, 1,5-anhydroglucitrol (1,5-AG), and continuous glucose monitoring (CGM).

Fructosamine measurement has been commercially available since the 1980s, and refers to the product formed by the nonenzymatic reaction of a sugar and a protein (in this case glucose and albumin). Because the half life of albumin is much shorter than that of an erythrocyte (about 20 days vs. about 120 days), fructosamine reflects a much shorter period of glycemic control compared to A1c, typically the preceding 2 to 3 weeks.^9,10^ Therefore, it provides a better assessment of recent changes in glycemic control.

However, fructosamine measurement also has limitations. Since the assay depends on albumin concentrations, patients with hypoproteinemia/hypoalbuminemia such as nephrotic syndrome or severe liver disease may have falsely low levels. It is unclear whether or not the assay should be corrected for serum protein or albumin concentrations. Other reported interferences include uremia, lipemia, and ascorbic acid.^10^ Because of these limitations, fructosamine is not considered a replacement for A1c. Its utility as a measure of glycemic control is reserved for when A1c may be inaccurate, or to assess recent changes in glycemic control.^11,12^

The term fructosamine encompasses all glycated proteins. Glycated albumin is an example of a fructosamine also used as a measure of glycemic control. Glycated albumin is typically reported as a percentage of total albumin, and also reflects glycemic control over the preceding 2 to 3 weeks. Its use has been particularly advocated in patients with end stage renal disease where A1c typically underestimates glycemic control, and glycated albumin would provide a more accurate assessment of recent glycemia.^13–15^

Plasma 1,5-anhydroglucitol (1,5-AG) is a naturally occurring dietary polyol that that has been proposed as a measure of glycemic control. Renal reabsorption of 1,5-AG is competitively inhibited by glucose. Hence, there is a negative correlation between 1,5-AG levels and glycemic control. Plasma 1,5-AG is a marker of short-term glycemic control, typically reflecting glucose values over the preceding 48 h to 2 weeks. The test may be useful to detect post-prandial hyperglycemia and glycemic variability, especially in the presence of target A1c. This test is affected by renal hemodynamics and may not be a reliable indicator of glycemia in renal failure (false high), pregnancy (false low), or chronic severe liver disease. In 2003, the FDA approved an automated, commercially available assay (GlycoMark™) to measure 1,5-AG levels.^16,17^ Some insurance plans may not cover Glycomark ™.

Another method used to reflect glycemia is continuous glucose monitoring (CGM). CGM sensors measure interstitial glucose levels on a semi-continuous basis, and reflect glycemic fluctuations while the device is worn. Three CGM devices are currently FDA approved for use in the United States (Medtronic CGM™, and Dexcom SEVEN PLUS™ and G4 PLATINUM™), and patients can wear their accompanying disposable sensors for 3, 7, and 7 day periods respectively. Data obtained from wearing a CGM sensor for up to 5 days has been shown to correlate well with A1c.^18^ CGM readings may be inaccurate during periods of high glycemic variability, as well as after ingestion of acetaminophen or ascorbic acid.^19,20^ Recent guidelines recommend clinicians consider using daily CGM in essentially all adult patients with type 1 diabetes, and intermittent CGM in any adult patient with diabetes and suspected nocturnal hypoglycemia, dawn phenomenon, postprandial hyperglycemia, hypoglycemic unawareness, or when significant therapeutic changes are made such as initiating or intensifying insulin.^21^ Insurance plans reimburse charges for insertion and interpretation, particularly in patients with type 1 diabetes and in patients with type 2 diabetes treated with insulin.

Although frucotsamine, glycated albumin, 1,5-AG, and CGM have been shown to correlate with mean glucose levels,^16^ it is important to remember that only A1c has been validated as a measure of both long-term glycemic control and risk for development of complications.^22^ Therefore, A1c remains the gold standard to measure average glycemic control. The other methods described should only be used in situations where A1c results may be inaccurate or misleading, or to supplement A1c data.

## Conclusion

Chronic liver disease affects glucose metabolism, ranging from mere glucose intolerance to overt diabetes, which is known as hepatogenous diabetes.We find that, about 50, 54% of chronic hepatitis C patients are diabetics with 25, 5% have normal HA1C, 74, 5% have abnormal levels. With no limitations, results precisely answer our question, demonstrate that hepatogenous diabetes is a common problem among chronic liver patients and HA1C is not a standard assessment tool for diabetes

### Recommendations

- More researches to explain the pathological basis of the mysterious relation between cirrhosis and HA1C.
- Comparative studies between different markers of diabetes
- Screening for diabetes among hepatitis patient on larger scale with better and more concise tools and under observation from a treating team of endocrinologists, hepatologists and clinical pathology

## Data Availability

All data produced in the present study are available upon reasonable request to the authors

## LIST OF ABBREVIATIONS

(VF-IFH): Virology unit at Ismailia fever hospital
(SCUH-Hep): Suez Canal university hospital hepatology wards

## Introduction

After meal, carbohydrates in the food we eat are reduced into simplest form, glucose. Excess glucose removed from body and converted into glycogen in a process called glycogenesis. 1

During fasting or when blood glucose level declines, hepatic cells convert glycogen into glucose and release them into blood till level of glucose approaches normal in a process called glycogenolysis. In chronic liver disease: there is progressive destruction of liver parenchyma over a period greater than 6 months leading to fibrosis and cirrhosis. Due to progressive loss of hepatocyte function, glucose metabolism is impaired in a process called hepatogenous diabetes.

What is hepatogenous diabetes? DM which develops as a complication of cirrhosis is known as Hepatogenous Diabetes

Patient with CLD may have two types of diabetes; type 2 DM: related to metabolic syndromes that cause non-alcoholic fatty liver disease, hepatocellular carcinoma. Hepatogenous Diabetes that result as a result of liver cirrhosis. Many studies approved that type 2 diabetes and hepatogenous diabetes are associated with increased risk of complication of chronic lver diseases and mortality.

Pathophysiology of hepatogenous diabetes is complex and not clearly understood. Hepatogenous diabetes may result from; Decrease extraction of insulin by damaged liver. And liver dysfunction itself has a toxic effect on pancreatic islets and this proved when hepatogenous diabetes improves after successful liver transplantation. As well as insulin Resistance of peripheral tissues (muscles, liver, fat) one of the major mechanism involved in pathophysiology. Genetic factors play a major role as well. HCV is considered a diabetogenic agent through multiple mechanisms: autoimmune phenomena, direct cytotoxic effect on pancreatic cells, and, blockage of insulin receptors at cellular levels. Alcoholic chronic liver disease affect both hepatocytes and pancreatic islet cells.

So in our study we want to identify best investigation to assess diabetes in chronic liver disease patients. Three prospective studies were collected assessing impact of diabetes among chronic liver disease patients; mainly the outcome, and all of them demonstrate lower 5-year cumulative survival

## Aim & Objectives

To provide data to augment the standard of care in diabetic patients with chronic liver disease

- OBJECTIVES
- Primary Objectives
  1. To determine prevalence of type2 diabetes mellitus among patients with chronic liver disease.
  2. To determine effect of chronic liver disease on credibility of glycosylated hemoglobin (Hb A1C) as a standard assessment tool for diabetes.
- Secondary Objectives
  1. To assess prevalence of family history among diabetic patients with chronic liver disease.
  2. To assess treatment modalities among diabetic patients with chronic liver disease.

## Subjects & Methods

- Data Collection Tools & procedures:
- Through a formal permission and access to the (VF-IFH) data and recording system.
- Data are recorded in a paper-based database system. Collection of data will be via copying the data into an excel sheet.
- - Diabetic status; fasting glucose will be used as a gold standard to divide patients into diabetics (abnormal fasting glucose) and non-diabetics (normal fasting glucose level).
- - Assessment of HbA1C values and patients’ diabetic control, as documented in (VF-IFH) database and records and by using … questionnaire

## Results

The sample size for this study is 167 of chronic hepatitis C patients. Out of 167 questioned patients, 30.54% are diabetics. 25.5% of diabetic patients have normal HA1C (controlled) & 74.5% have abnormal HA1C (uncontrolled) 78.43% of patient have elevated fasting plasma glucose & about 21.57% have normal values. About 56.86% of hepatitis C patients that have diabetes, have abnormal kidney function (elevated serum creatinin)

## Conclusion

- Chronic liver disease affects glucose metabolism, ranging from mere glucose intolerance to overt diabetes, which is known as hepatogenous diabetes.
- We find that, about 50, 54% of chronic hepatitis C patients are diabetics with 25, 5% have normal HA1C, 74, 5% have abnormal levels.
- With no limitations, results precisely answer our question, demonstrate that hepatogenous diabetes is a common problem among chronic liver patients and HA1C is not a standard assessment tool for diabetes.

## Appendix

**Clinical questionnaire assessing diabetic progression**

**Name**_______________ Date__________________

1. **Are you (check one)** Male Female
2. How old a**re you?**____________ years old
3. **when you were first diagnosed with diabetes ?** year ……… Age…..
4. **Has your doctor prescribed pills for your diabetes?** ____________ Yes ____________ No
5. **How often are you supposed to take these pills?** ____________ I do not take pills for my diabetes ____________ Occasionally as needed ____________ Once per day ____________ Twice per day ____________ Three or more times per day
6. **Has your doctor prescribed insulin shots for your diabetes?** ____________ Yes ____________ No
7. **How often are you supposed to take insulin?** ____________ I don’t take insulin ____________ Occasionally as needed ____________ Once a day ____________ Twice a day ____________ Three or more times a day
8. **Do you follow your prescribed medication reguraly ?** ____________ yes ____________ No
9. **How often do you test your blood glucose?** ____________ Occasionally as needed ____________ A couple times a month ____________ 1 or 2 times a week ____________ 3 to 6 times a week ____________ Once a day
10. 10 **-In general, would you say your health is:** Excellent ………………………… Very good……………………….. Good……………………………….. Fair…………………………………. Poor…………………………………
11. **Have you had low blood sugar reactions lately?** ____________ Yes ____________ No
12. **Have you had symptoms of high blood sugar lately?** ____________ Yes ____________ No **Which?** (fatigue, hunger, thirst, frequent desire to urinate, trouble focusing vision)
13. **Have you had problems with infections?** ____________ Yes ____________ No Which? (acne, burning on urination, frequent colds, itching in groin or feet, boils)
14. **Have you ever been told you have High Blood Pressure?** Yes No if yes. on medication or not ?
15. **Have you ever had Problems with your eyes ?** Yes No if yes. what ?
16. **When was the last time you had an eye exam in which the pupils were dilated? This would have made you temporarily sensitive to bright light**. LESS THAN 1 MONTH …………………………… 1-12 MONTHS……………………………………….. 13-24 MONTHS……………………………………… GREATER THAN 2 YEARS …………………….. NEVER ………………………………………………….
17. **Has a doctor ever told you that diabetes has affected your eyes or that you had retinopathy?** YES ……………………………………………………… NO ………………………………………………………..
18. **Have you ever had your cholesterol tested?** Yes No If YES, How long ago?
19. **Have you ever been diagnosed with elevated cholesterol?** Yes No
20. **If you have been diagnosed with elevated cholesterol, are you on treatment?** Yes No
21. **Do you have lung disease?** Yes No If YES, What? ______________________________
22. **Have you ever had heart disease?** Yes No, If YES, what? _____________________________
23. **Have you ever had a heart catheterization?** Yes No If YES, when? ______________________________
24. **Have you ever had a heart attack?** Yes No If YES, when?_____________________
25. **have you ever had Numbness or tingling in your feet or legs ?** Yes No
26. **do you have any amputed limbs ?** Yes No if yes cause ?
27. **In the past month, rating 0 as no problem and 10 as having a severe problem please rate**

**…………**

## REFERENCES

1. Nordlie RC, Foster JD, Lange AJ. Regulation of glucose production by the liver. Annu Rev Nutr. 1999;19:379–406. doi:10.1146/annurev.nutr.19.1.379.

2. Garcia-Compean D. Liver cirrhosis and diabetes: Risk factors, pathophysiology, clinical implications and management. World J Gastroenterol. 2009;15(3):280. doi:10.3748/wjg.15.280.

3. García-Compeán D, González-González JA, Lavalle-González FJ, González-Moreno EI, Villarreal-Pérez JZ, Maldonado-Garza HJ. Hepatogenous diabetes: Is it a neglected condition in chronic liver disease? World J Gastroenterol. 2016;22(10):2869. doi:10.3748/wjg.v22.i10.2869.

4. Holstein A, Hinze S, Thiessen E, Plaschke A, Egberts E-H. Clinical implications of hepatogenous diabetes in liver cirrhosis. J Gastroenterol Hepatol. 2002;17(6):677–681. http://www.ncbi.nlm.nih.gov/pubmed/12100613.

5. Nishida T, Tsuji S, Tsujii M, et al. Oral glucose tolerance test predicts prognosis of patients with liver cirrhosis. Am J Gastroenterol. 2006;101(1):70–75. doi:10.1111/j.1572-0241.2005.00307.x.

6. Keohane EM, Walenga JM, Smith LJ, Rodak BF. Rodak’s Hematology: Clinical Principles and Applications.

7. Bunn HF, Haney DN, Kamin S, Gabbay KH, Gallop PM. The biosynthesis of human hemoglobin A1c. Slow glycosylation of hemoglobin in vivo. J Clin Invest. 1976;57(6):1652–1659. doi:10.1172/JCI108436.

8. Saudek CD, Brick JC. The Clinical Use of Hemoglobin A1c. J Diabetes Sci Technol. 2009;3(4):629–634. doi:10.1177/193229680900300402.

9. Tahara Y, Shima K. Kinetics of HbA1c, Glycated Albumin, and Fructosamine and Analysis of Their Weight Functions Against Preceding Plasma Glucose Level. Diabetes Care. 1995;18(4):440–447. doi:10.2337/diacare.18.4.440.

10. Nitin S. HbA1c and factors other than diabetes mellitus affecting it. Singapore Med J. 2010;51(8):616–622. http://www.ncbi.nlm.nih.gov/pubmed/20848057.

11. Kruszynska YT, Home PD, McIntyre N. Relationship between insulin sensitivity, insulin secretion and glucose tolerance in cirrhosis. Hepatology. 1991;14(1):103–111. doi:10.1002/hep.1840140117.

12. Megyesi C, Samols E, Marks V. GLUCOSE TOLERANCE AND DIABETES IN CHRONIC LIVER DISEASE. Lancet. 1967;290(7525):1051–1056. doi:10.1016/S0140-6736(67)90334-0.

13. Koga M, Kasayama S, Kanehara H, Bando Y. CLD (chronic liver diseases)-HbA1C as a suitable indicator for esti

